# Optimising the implementation of a universal web-based mental health service for Australian secondary schools: A cluster randomised controlled trial

**DOI:** 10.1101/2025.02.14.25322324

**Authors:** Mirjana Subotic-Kerry, Andrew Mackinnon, Dervla Gallen, Simon Baker, Belinda Louise Parker, Melinda Rose Achilles, Cassandra Chakouch, Nicole Cockayne, Helen Christensen, Bridianne O’Dea

## Abstract

**Background:** Secondary schools are increasingly delivering a range of mental health interventions with varied success. This trial examined the effectiveness of two implementation strategies, class time allocation, and financial incentives, on secondary students’ engagement with a universal web-based mental health service, ‘Smooth Sailing’.

**Methods:** A three-arm, cluster-randomised trial was conducted over 12 weeks with Grade 8 and 9 students from 20 schools in two Australian states. The primary outcome was student engagement, measured by the number of modules accessed at 12-weeks post-baseline. Secondary outcomes included uptake, retention, help-seeking intentions for mental health problems, service satisfaction, and barriers to use.

**Results:** A total of 20 schools consented to take part and 1295 students consented to the study. The mean number of modules accessed by students was higher in the enhanced conditions (class time allocation and financial incentive) but not statistically significant (P>0.140). There were no significant differences in uptake (P=0.554) or retention (P=0.945) between conditions. Help-seeking intentions significantly improved at 6- and 12-weeks in the standard service and class time conditions only. Common barriers to service use among students were forgetfulness and low motivation.

**Conclusions:** None of the implementation strategies examined by this trial significantly increased student engagement when measured by modules accessed.

**Trial registration:** Australian New Zealand Clinical Trial Registry (ACTRN12621000225819) and Universal Trial Number (U1111-1265-7440).

**Trial funding:** The trial operations were funded by the Prevention Hub, Australian Government Department of Health.

## INTRODUCTION

Digital mental health interventions are increasingly being delivered in schools to improve access to mental health resources and support student well-being (e.g., Bevan Jones et al., 2023). Secondary schools, particularly in Australia and the United States, have become key settings for delivering mental health interventions, with students utilising school-based services as often as they do in traditional healthcare settings (Duong et al., 2021; Lawrence et al., 2015). However, schools, like other service sectors, face significant implementation challenges that limit the uptake, integration, and sustainability of evidence-based interventions. While schools are diverse and dynamic settings, few studies have gone beyond the confines of clinical trials to examine the ‘real-world’ translation of these interventions (Hagermoser Sanetti & Collier-Meek, 2019). Consequently, many school-based mental health interventions have failed to achieve scalability. Evaluating the effectiveness of implementation strategies to increase service engagement among students may help to increase the reach and impact of school-based mental health interventions.

School-based mental health programs and services vary in intervention type, delivery method, support levels, and youth involvement. Universal interventions, which target all students regardless of their mental health status or risk, offer several advantages over targeted approaches. They increase protective factors, reduce stigma (Greenberg & Abenavoli, 2017; Werner-Seidler et al., 2017), and align with whole-of-school well-being policies, making them a preferred option by schools (Beames et al., 2021; Horowitz & Garber, 2006). Universal, digital mental health programs have been shown to improve emotional outcomes such as resilience, coping skills, and self-efficacy in young people (Fenwick-Smith et al., 2018). Small improvements in help-seeking intentions (O’Dea, Subotic-Kerry, et al., 2021), and modest effects on depression and anxiety outcomes have also been reported (Hugh-Jones et al., 2021; Werner-Seidler et al., 2021). However, low engagement and high dropout rates have been a major issue in past interventions (Badawi et al., 2023), and significant challenges remain in optimising student engagement and adherence to these programs (Bevan Jones et al., 2023; Bevan Jones et al., 2020; Proctor et al., 2009).

Improving student engagement with digital mental health programs is crucial for ensuring adequate exposure to the therapeutic content within school-based interventions. A systematic review and meta-analysis found significant associations, albeit modest, between engagement and improved mental health outcomes (effect sizes: r=0.23 to Hedges′ g=−0.40), with module completions emerging as a key engagement metric (Gan et al., 2021). Similarly, Donkin et al. (2011), found that improvements in individuals’ mental health outcomes were linked to the number of modules accessed, but not to other engagement metrics such as time spent or logins. Strategies such as allocating class time and teacher support may significantly enhance completion rates when compared to self-directed completions. For example, school-based interventions supervised by teachers have been found to have higher completion rates than those accessed independently by students in the community (Neil et al., 2009). Additionally, human guidance, including feedback and support, has been shown to increase completion rates in web-based interventions for depression and anxiety in adults (Musiat et al., 2022).

Research suggests that financial incentives may also enhance engagement with digital programs. A recent review found that app-based programs offering monetary incentives had higher retention rates compared to those without incentives (Anguera et al., 2016; Pratap et al., 2020). Similarly, in a randomised controlled trial, adolescents were more likely to complete an online, self-guided, single-session mental health intervention when paid, compared to those in an unpaid, naturalistic setting (Cohen & Schleider, 2022). Small financial incentives have also been shown to improve adherence to a web-based intervention promoting physical activity in adults (Wurst et al., 2020), and increased survey response rates and patient compliance (Sauermann & Roach, 2013).

### Implementation of the Smooth Sailing service

The Smooth Sailing service is a universal, web-based mental health service for secondary school students developed by the Black Dog Institute (O’Dea et al., 2019; O’Dea, Subotic-Kerry, et al., 2021). As shown in Figure 1, the service screens students’ symptoms of anxiety and depression using validated self-reported scales and then offers self-directed psychoeducation and internet-delivered Cognitive Behavioural Therapy (iCBT) modules for students with nil to moderate symptoms (Buttazzoni et al., 2021; Christ et al., 2020), and in-person support from school counsellors for those with severe symptoms (O’Dea et al., 2019; O’Dea, Subotic-Kerry, et al., 2021). A randomised controlled trial showed that the service significantly improved help-seeking intentions and anxiety symptoms in secondary students (O’Dea, Subotic-Kerry, et al., 2021), with minimal potential for harms (Braund et al., 2024). However, module completion was low. Students reported time, low motivation, and the perceived lack of need for care as barriers to engagement (O’Dea, Subotic-Kerry, et al., 2021). To overcome this, school counsellors suggested embedding rewards or incentives and providing students with dedicated class time to complete the modules (O’Dea, King, et al., 2021). However, little is known about the effectiveness of these approaches.

**Figure 1.**
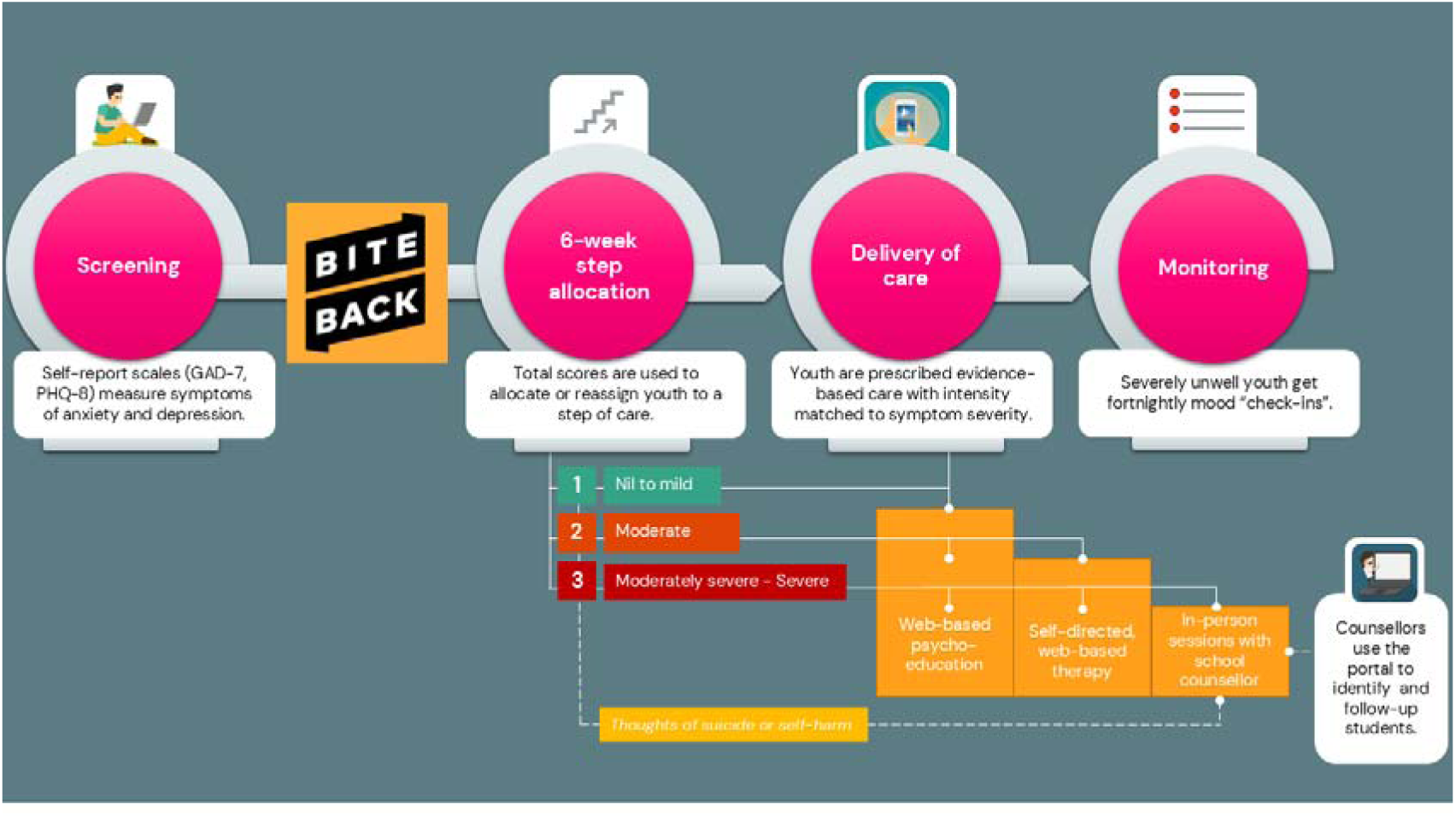
The revised Smooth Sailing service model: Step criteria and care provided.

The School Implementation Strategies Translating ERIC Resources (SISTER) framework (Cook et al., 2019) was used to modify Smooth Sailing with evidence-based strategies (see Supplementary Materials, Table 1). To increase the engagement of non-symptomatic students in the service, a 6-week “watchful waiting” period was introduced (see Figure 1). During this period, all participating students received the Bite Back Six-Week Challenge, a curriculum-aligned, positive psychology program (Burckhardt et al., 2015; Manicavasagar et al., 2014). To address the perceived lack of need for care among students, we revised the student feedback mechanism to include a detailed report on their symptoms, including age norms, functional impacts, and reasons for engaging with the service. To increase the accessibility of the interventions, an evidence-based smartphone app was integrated into the iCBT module, complementing the existing web-based programs (O’Dea et al., 2020).

**Table 1.** Participant characteristics at baseline stratified by condition (N=1295)

|  | SS<br>(n=469) |  | SS-CT<br>(n=335) |  | SS-FI<br>(n=491) |  | Total sample<br>(N=1295) |  |
| --- | --- | --- | --- | --- | --- | --- | --- | --- |
|  | M | SD | M | SD | M | SD | M | SD |
| Age (years) | 13.72 | 0.70 | 13.77 | 0.74 | 14.30 | 0.61 | 13.95 | 0.73 |
| Help-seeking intentions<br>(GHSQ) | 33.55 | 10.74 | 33.58 | 10.44 | 34.14 | 9.76 | 33.79 | 10.30 |
|  | *n* | % | *n* | % | *n* | % | *n* | % |
| Female | 232 | 49.5 | 121 | 36.1 | 290 | 59.1 | 643 | 49.7 |
| Sexuality diverse<sup>a</sup> | 42 | 9.0 | 25 | 7.5 | 31 | 6.3 | 98 | 7.6 |
| Aboriginal or Torres Strait<br>Islander | 40 | 8.5 | 33 | 9.9 | 45 | 9.2 | 118 | 9.1 |
| Previous or current mental<br>health problem or diagnosis<br>of a mental illness | 129 | 27.5 | 106 | 31.6 | 107 | 21.8 | 342 | 26.4 |
| Currently receiving<br>psychological therapy | 35 | 7.5 | 47 | 14.0 | 41 | 8.4 | 123 | 9.5 |
| Currently taking<br>medication for a mental<br>illness | 17 | 3.6 | 26 | 7.8 | 13 | 2.6 | 56 | 4.3 |
| Previously or currently had<br>a session with a school<br>counsellor | 122 | 26.0 | 92 | 27.5 | 138 | 28.1 | 352 | 27.2 |

Note: <sup>a</sup> Identifies as either lesbian, gay, bisexual, transgender, queer, or intersex, LGBTQI.

### Objectives of the current trial

This study examined the effectiveness of two student-level implementation strategies, (class time allocation, financial incentives), on secondary school students’ uptake, engagement, and retention in the Smooth Sailing service. It was hypothesised that these strategies would increase student engagement, measured by the number of modules accessed, compared to the standard service. Service satisfaction and barriers to use were also assessed. Additionally, this study explored the impact of these implementation strategies on the primary outcome of the service (help-seeking intentions for mental health problems). It was hypothesised that increased engagement would mediate improvements in help-seeking intentions.

## METHODS

### Design

A three-arm, cluster randomised controlled trial (RCT) was conducted with schools as clusters and students as individual participants. The reporting adheres to the CONSORT checklist for RCTs (Cuschieri, 2019). All outcome measures were assessed at the individual level. Data collection occurred at baseline, 6-weeks post-baseline, and 12-weeks post-baseline, with the primary outcome evaluated at 12-weeks post-baseline (primary endpoint). All participants reached the primary endpoint by December 2020. Ethical approval was granted by the University of New South Wales Human Research Ethics Committee (HC190382), the New South Wales Department of Education State Education Research Applications Process (2019302), the Catholic Schools Offices for the Dioceses of Maitland-Newcastle, Parramatta, and Wagga Wagga. The trial was registered with the Australian New Zealand Clinical Trials Registry (ACTRN12621000225819) and assigned the Universal Trial Number (U1111-1265-7440).

### Participants

The trial was conducted in secondary schools across South Australia and New South Wales, Australia. Schools were eligible to participate if they provided written Principal consent, had an onsite school counsellor or equivalent, and had reliable Internet access. All Grade 8 or 9 students with an active email address were eligible to participate unless their parent(s)/guardian(s) opted them out.

### Sample size

The target sample size was 1350 students (approximately 450 per condition), calculated to detect a small effect size (.30) on a continuous primary outcome, with an alpha of 0.5, and a 20% attrition rate. A conservative intraclass correlation (ICC) of 0.02 was assumed for possible clustering effects, slightly higher than the previous ICC of 0.017. Based on previous data (O’Dea, Subotic-Kerry, et al., 2021), approximately 85 students per school were expected, requiring a minimum of 15 schools to meet the target sample size.

### Randomisation and blinding

Cluster randomisation at the school level was used to prevent contamination and for administrative feasibility (Flynn, 2001). Once principal consent was obtained, randomisation was performed by an external researcher not involved in the day-to-day trial activities, following International Council for Harmonisation (ICH) guidelines (Lewis, 1999). Schools were allocated in a 1:1 ratio using a minimisation approach (Altman & Bland, 2005; Taves, 2010) to balance conditions by the Index of Community Socio-Educational Advantage (ICSEA) level (<1,000; >1,000), school type (co-educational; gender-selective), and grade level (Grade 8; Grade 9). Minimisation was conducted in Stata version 14.2 using the rct_minim procedure (Ryan, 2018).

The initially available pool of schools was randomly sorted using Microsoft Excel random number generator, and schools entered the minimisation routine in ascending order using the balancing variables. Schools that consented later were allocated using the same minimisation procedure as they provided complete information. While the research program manager and one research officer were aware of the allocations due to administrative requirements, the broader research team remained blinded until the first school visit. Schools were informed of their assigned condition via email after they had consented. Students were not informed of their allocations, except for those in the financial incentive condition, who were notified at the 6-week mark when the first incentive became available.

### Recruitment and consent

Schools were recruited via direct email invitations sent to relevant staff from the Black Dog Institute’s national school database. Additional recruitment efforts included advertisements in the Institute’s NSW School Counsellors e-Newsletter, the NSW School-Link newsletter (a government initiative that links schools with metropolitan and regional health services), the Black Dog Institute website, and social media platforms (Facebook and Twitter), academic conferences, professional development courses, and state Departments of Education.

Interested schools contacted the research program manager, who provided a study information package and followed up by phone. Once the school principal and school counsellor submitted a signed consent form, and schools were allocated to their study condition, the research team scheduled the school visits. Two weeks before the first visit, schools were instructed to distribute student information forms and a brief video about the service, with schools circulating service information through newsletters and other channels. Each participating school received a comprehensive implementation guide that outlined the service’s purpose along with clear, step-by-step instructions for setting up the service. This included guidance on school registration, forming a delivery team, selecting year groups, and promoting the service. The guide also detailed the screening process at 6- and 12-weeks, follow-up notifications for school counsellors, and how to use the School Counsellor Portal to ensure student safety. A supplementary guide was provided to School Counsellors with more detailed information about the School Counsellor Portal, and schools assigned to provide additional class time sessions were given a supplementary guide with further information about arranging the extra class time sessions.

An opt-out consent process was used, allowing parents to decline their child’s participation by notifying the school or research team before the first visit. In the Wagga Wagga Catholic Diocese, parental/guardian consent was required via an online or hardcopy form before the first visit. For all other students, informed consent was provided online on the day of the first assessment. All students participating completed a five-item Gillick competence test (Kelly & Halford, 2007) to ensure student understanding of the study.

### Procedure

The study procedure was consistent across the three conditions, except students in the class time condition who received extra class time to use the Smooth Sailing service, and students in the financial incentive condition who received service reminders with tailored content. The research team conducted three assessment sessions during class time. Initially intended as face-to-face, these sessions were adapted to teleconferencing due to COVID-19, allowing schools to choose their preferred format.

At the first assessment (baseline), students accessed the Smooth Sailing website via a school-specific URL, completed informed consent, registered by providing their name, gender, age, email, and mobile number (optional), and created a username and password.

Mental health screening was conducted at baseline and repeated at 6- and 12-weeks post-baseline through a URL sent to students’ registered email (see Supplementary Material for more detail).

After each assessment, researchers met with school counsellors either in-person or online to ensure they could access and navigate the web portal for student follow-ups. School counsellors were provided with guides to interpret mental health scores and were instructed to follow school protocols for student support, referrals, and parental contact when needed. They were also given a list of local mental health services. Researchers used standard instruction sheets for these meetings and did not provide clinical supervision. Three days after each school visit, researchers reviewed the School Counsellor Portal to ensure all follow-ups were completed and monitored for adverse events. If necessary, they contacted school counsellors to update the portal accordingly.

### Implementation strategies and conditions

#### Control condition – Standard service (SS)

Schools and students received the standard Smooth Sailing service. Screening activities occurred during class and students completed the service modules independently, in their own time.

#### Standard service + class time (SS-CT)

In addition to the standard service, students in this condition received two extra class periods to access the service and complete modules. There were no restrictions on the type or duration of the class periods; however, most Australian secondary school class periods range from 45–75 minutes.

#### Standard service + financial incentive (SS-FI)

In addition to the standard service, students in this condition were informed at the 6-week mark that they would be reimbursed 3.0AUD for each of the first five modules completed and 2.5AUD each for the 6- and 12-week screening assessments, with a maximum reimbursement of 20.0AUD.

### Outcome measures

#### Service engagement

The primary outcome was the number of modules accessed by students. The Smooth Sailing service included seven modules: Bite Back Six-Week Challenge, five psycho-education modules, and one iCBT module. Engagement was measured as the proportion of modules accessed, which varied based on students’ step allocation at 6- and 12-weeks.

#### Service uptake

Uptake was measured by the number of students who registered for the service at baseline.

#### Service retention

Retention was measured by the proportion of registered students who completed the 12-week mental health screener.

#### Help-seeking intentions for mental health problems

The likelihood of seeking help for mental health problems was assessed using the General Help-Seeking Questionnaire (GHSQ: Wilson et al., 2005) at baseline, 6- and 12-weeks post-baseline. Responses were rated on a 5-point Likert scale from “extremely unlikely (1)” to “extremely likely (5)” (α=.89). Higher scores indicated a greater likelihood of seeking help from 13 sources, with an option to specify additional sources. The GHSQ is widely used among adolescents aged 11-18 years and has shown satisfactory psychometric reliability (Divin et al., 2018).

#### Service satisfaction and barriers to use

Service satisfaction was evaluated at 12-weeks post-baseline using an 11-item questionnaire assessing enjoyment, ease of use, usefulness, and comfort. Participants rated agreement with statements about the service and provided an overall helpfulness rating on a 5-point Likert scale. Barriers to use were evaluated with an 18-item questionnaire assessing technical, personal, and intervention-specific barriers, with responses recorded as “yes” or “no”.

#### Demographics and mental health history

Students provided their age, gender, sexual identification, and Aboriginal and Torres Strait Islander status. They were asked whether they had ever experienced or been diagnosed with a mental health illness such as depression or anxiety (yes, no, I’d rather not say). Additionally, students indicated if they were currently receiving treatment or taking prescribed medication for a mental health condition using the same response options. A separate question asked whether they had ever attended a session with the school counsellor (yes, no, I’d rather not say).

### Data collection and analysis

Data were securely collected and stored using the Black Dog Institute research engine hosted on the University of New South Wales servers. Data was extracted using Microsoft Excel and analysed using SPSS version 27.0 (SPSS Inc, Chicago, I1, USA, 2017). Analyses followed an intention-to-treat approach, including all randomised participants regardless of the intervention received.

To examine module access, a series of models for count data were fitted to the number of modules accessed by each participant, with group assignment included as a fixed factor and school as a random factor to account for clustering effects. Based on the 6- and 12-week screening assessments after Bite Back completion, the maximum number of available modules per participant was included as an exposure offset variable. The cluster-specific incident rate ratio (IRR) was used to compare module completion rates between each enhanced condition and the standard service for an ‘average’ school (i.e., a school with a random effect of 0). Individual school effects and the model’s overdispersion parameter were considered.

To evaluate the population-level impact of each intervention on module access in the full sample, the intraclass correlation coefficient (ICC) was estimated using the trigamma function to assess observation-level variance, including the maximum number of modules as a fixed effect (Nakagawa, 2017). Given the limitations of count and logistic models in accommodating the increased proportion of participants accessing more modules at the upper distribution, alternative model formulations such as the proportional odds model were explored. This model assumes constant odds of transitioning between modules regardless of the number of modules accessed.

Uptake and retention rates were analysed using mixed-effects logistic regression, with school included as a random effect to accommodate for clustering effects. The effects of the implementation strategies on help-seeking intentions were assessed using linear mixed-effects models, comparing changes over time and differences between conditions. A sub-group analysis was also conducted to explore the relationship between service engagement and help-seeking intentions.

## RESULTS

### Sample

As shown in Figure 2, 119 schools expressed interest in the study and 26 (21.9%) agreed to participate. Nine schools were allocated to receive the standard service (*n*=5, 62.5% located in major cities), 7 to the additional class time condition (*n*=3, 42.9% located in major cities), and 10 were allocated to the incentive condition (*n*=6, 60% located in major cities). Two schools withdrew due to COVID-19 after allocation but before the baseline assessment.

**Figure 2.**
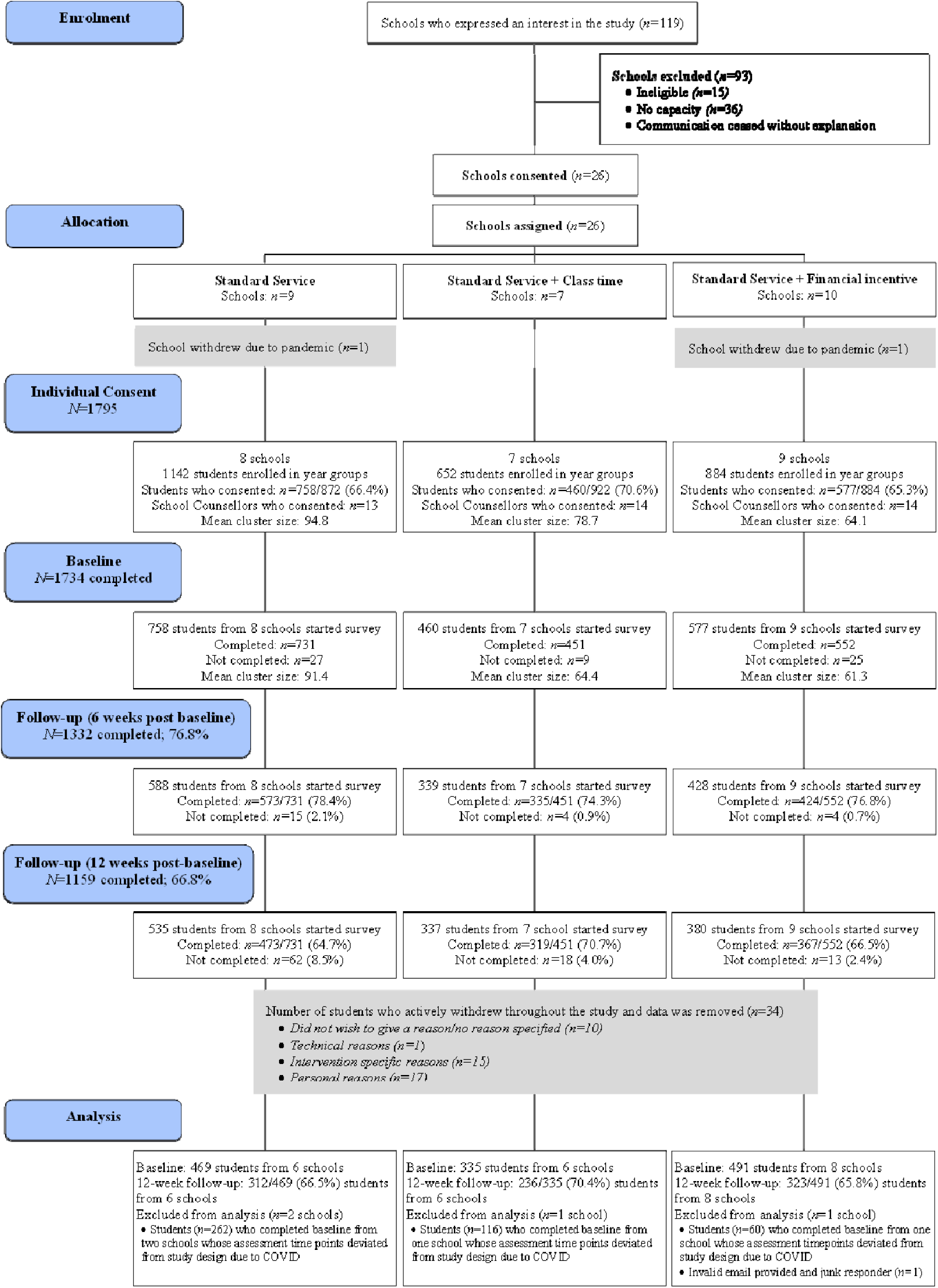
CONSORT dh1gnun showing participation, withdrawals, and attrition.

In total, 1795 students from 24 schools consented and completed the baseline assessment. However, due to COVID-19 closures, four schools started the trial in Term 1 between 11^th^ February 2020 and 19^th^ March 2020, but experienced interruptions and resumed data collection later in the year (23^rd^ July 2020 to 5^th^ August 2020), with the final school visits occurring in Term 3 (3^rd^ September 2020 to 17^th^ September 2020). The remaining 20 schools commenced the trial in Term 3, 2020 (10^th^ August 2020 to 17^th^ September 2020), and completed the trial in Term 4 2020 (2^nd^ November 2020 to 10^th^ December 2020). The four schools (*n*=438) that started in Term 1 were excluded from the analyses due to the extended delay between the first two study assessments. The final sample consisted of 1295 students from 20 schools.

In the final sample, half of the participants were female (*n=*643/1295; 49.7%), with a mean age of 13.95 years (SD: 0.73; range 12-16 years). One in four reported a past or current mental health problem (*n*=342/1295, 26.4%), 9.3% (*n*=123/1295) were receiving psychological therapy, and 27.2% (*n*=352/1295) had previously had a session or were currently seeing their school counsellor. Participant characteristics are outlined in Table 1.

### Service engagement: Modules accessed

Table 2 shows the number of students who accessed Bite Back and each module in the service.

**Table 2.** Total number of modules accessed by the total sample and stratified by condition (N=1295)

|  | SS<br>(n=469) |  | SS-CT<br>(n=335) |  | SS-FI<br>(n=491) |  | Total sample<br>(N=1295) |  |
| --- | --- | --- | --- | --- | --- | --- | --- | --- |
|  | *n* | % | *n* | % | *n* | % | *n* | % |
| Bite Back | 231 | 49.3 | 259 | 77.3 | 284 | 57.8 | 774 | 59.8 |
| Module 1 | 80 | 17.1 | 81 | 24.2 | 162 | 33.0 | 323 | 24.9 |
| Module 2 | 161 | 34.3 | 165 | 49.3 | 226 | 46.0 | 552 | 42.6 |
| Module 3 | 86 | 18.3 | 79 | 23.6 | 152 | 31.0 | 317 | 24.5 |
| Module 4 | 60 | 12.8 | 61 | 18.2 | 134 | 27.3 | 255 | 19.7 |
| Module 5 | 76 | 16.2 | 71 | 21.2 | 142 | 28.9 | 289 | 22.3 |
| Module 6<sup>a</sup> | 50 | 29.4 | 58 | 45.7 | 96 | 50.3 | 204 | 41.8 |

*Notes:* <sup>a</sup> Module 6 was only available to participants at Steps 2+ so the percentage was calculated by dividing the number of students who accessed this module by the number of users allocated to Steps 2+ at 6- or 12-weeks (*n*=170, 127, and 191 respectively).

As shown in Figure 3, a considerable proportion of participants across all conditions did not access any other modules or progress beyond the first module. Rates of module access also varied substantially between schools. Across all schools, 46.7% of students (*n*=605/1295) did not access Bite Back or any other modules. Over half of the students (54.8%, *n*=257/469) allocated to the standard service did not access any modules including

**Figure 3.**
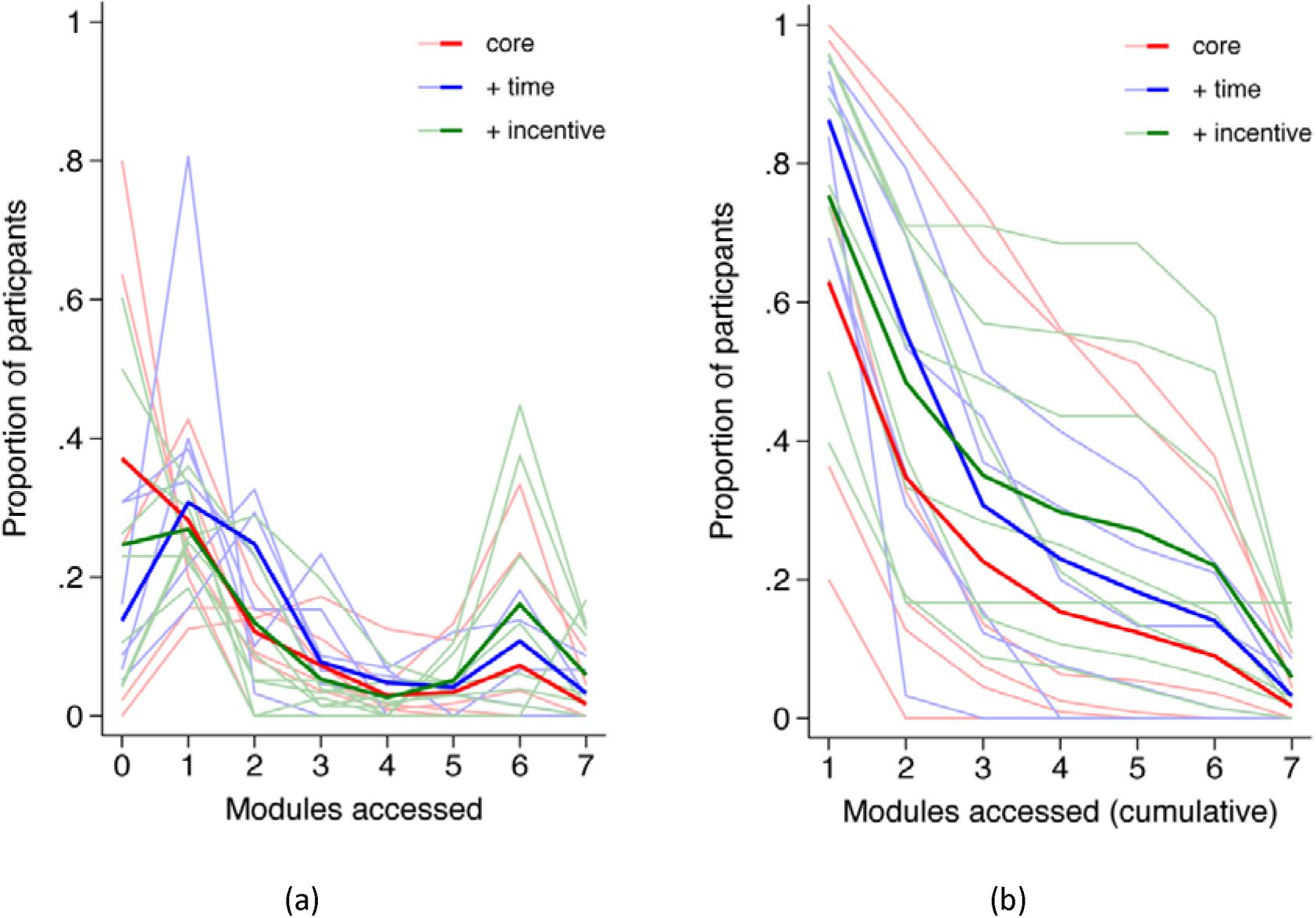
(a) Observed proportion and (b) cumulative proportion of modules accessed for each school (thin lines) and mean proportions (thick lines) for each condition.

Bite Back. Among students allocated to the class time condition, 38.8% (*n*=130/335) of students did not access any modules, while 44.4% (*n*=218/491) of students in the financial incentive condition did not access Bite Back or any other modules.

Table 3 shows the estimated mean count for each condition. While each implementation strategy increased the mean number of modules accessed, the improvement from the standard service was not statistically significant in either case (P>0.1404).

**Table 3.**
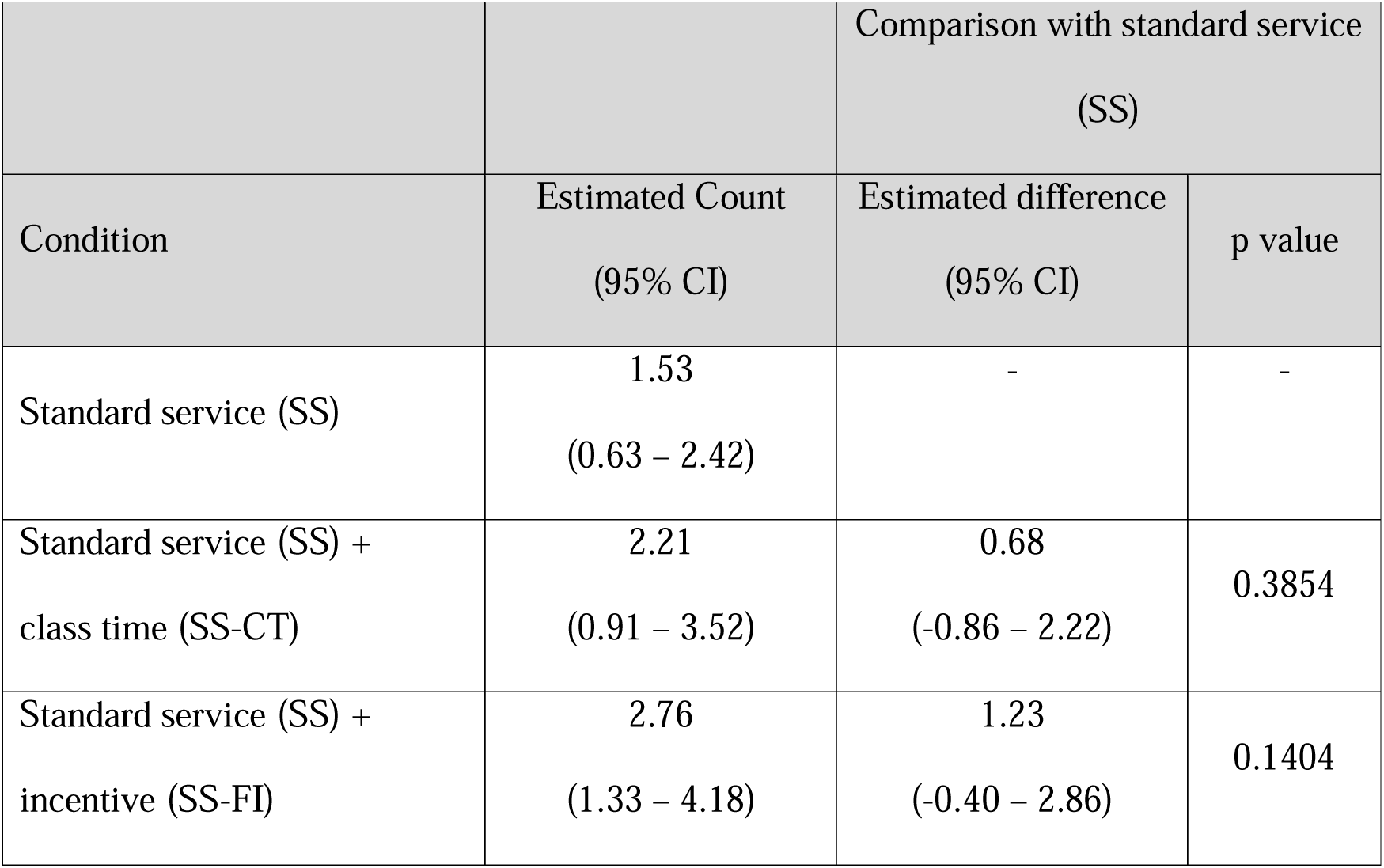
Estimated marginal mean number of modules accessed for each intervention and differences from standard service for the full sample.

| Condition | Estimated Count<br>(95% CI) | Comparison with standard service<br>(SS) |  |
| --- | --- | --- | --- |
|  |  | Estimated difference<br>(95% CI) | p value |
| Standard service (SS) | 1.53<br>(0.63 – 2.42) | - | - |
| Standard service (SS) +<br>class time (SS-CT) | 2.21<br>(0.91 – 3.52) | 0.68<br>(-0.86 – 2.22) | 0.3854 |
| Standard service (SS) +<br>incentive (SS-FI) | 2.76<br>(1.33 – 4.18) | 1.23<br>(-0.40 – 2.86) | 0.1404 |

A mixed effects negative binomial model fitted the data substantially better than a Poisson model (χ^2^ = 118.21, df=1, P<0.0001) and better than a negative binomial model omitting the random effect of school (χ^2^ =409.13, df=1, P<0.0001). Comparison of observed and expected counts and examination of Pearson residuals showed this model fitted the highly prevalent low modules accessed end of the data distribution extremely well, with the only source of poor fit attributable to high module access (6 and 7 modules), notably in the incentive condition. For the class time condition, the IRR of 1.45 (95% CI: 0.64 – 3.25) was not significant (z=0.90, P=0.3706). Similarly, the IRR of 1.80 (95% CI: 0.85-3.84) for the incentive condition was not significant *z*= 1.53, P=0.1263).

### Uptake and service retention

Figure 2 displays the uptake rates in the three conditions. Uptake rates for the standard service, class time, and financial incentive conditions for the final sample of students across the 20 schools were 59.1% (*n*=492/832), 65.1% (*n*=343/527), and 65.6% (*n*=514/784), respectively, with no significant differences between them (χ^2^=1.18, df=2, p=0.5545). Retention rates across the standard service, class time, and financial incentive conditions at 12-weeks post-baseline were 66.5% (*n*=312/469), 70.4% (*n*=236/335), and 65.8% (*n*=323/491) respectively, with no significant differences across the three conditions (χ^2^=0.11, df=2, p=0.9448).

### Effects of modules accessed on help-seeking intentions

There were no significant differences in help-seeking intentions between conditions at any time point. However, as shown in Figure 4, both the standard service and class time conditions showed significant improvements from baseline to 6-weeks post-baseline, with mean increases of 1.51 (i=3.30, df=1001.1, *P*=0.0010) for the standard service and 1.36 (*t*=2.42, df=1115.2, P=0.0158) for the class time condition. Similarly, significant improvements were observed at 12-weeks post-baseline, with mean increases of 1.16 (*t*=2.34, df=14.15.7, *p*=0.0196) for the standard service and 1.81 (*t*=3.14, df=1281.7, *p*=0.0017) for the class time condition. No significant changes were observed for the incentive condition at either time point (all P>.05).

**Figure 4.**
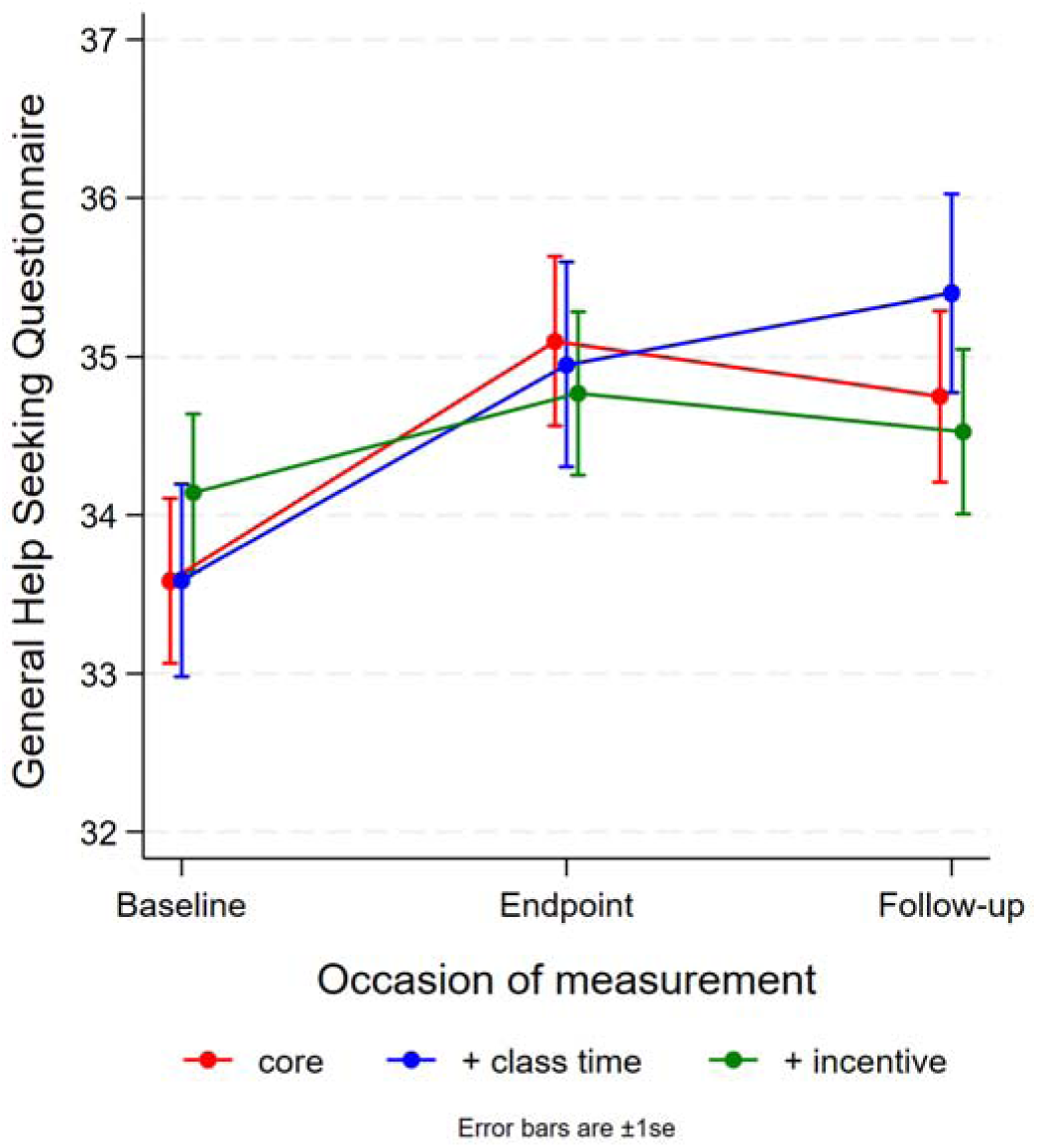
Help-seeking intentions by condition and occasion of measurement.

To explore the relationship between service engagement and help-seeking intentions, a categorisation of “no modules” versus “one or more modules” accessed was used. There were some significant differences in each category. As shown in Figure 5, among students who completed no modules, the incentive condition was associated with a significantly lower change in help-seeking intentions from baseline compared to students receiving the standard service. Conversely, students in the standard service and class time conditions showed improvement, while students in the incentive condition did not. For students who completed some modules, those in the incentive condition maintained relatively stable help-seeking intentions, initially starting slightly higher than those in the standard service and class time conditions. However, students allocated to receive the standard service and who were allocated additional class time caught up to varying extents.

**Figure 5.**
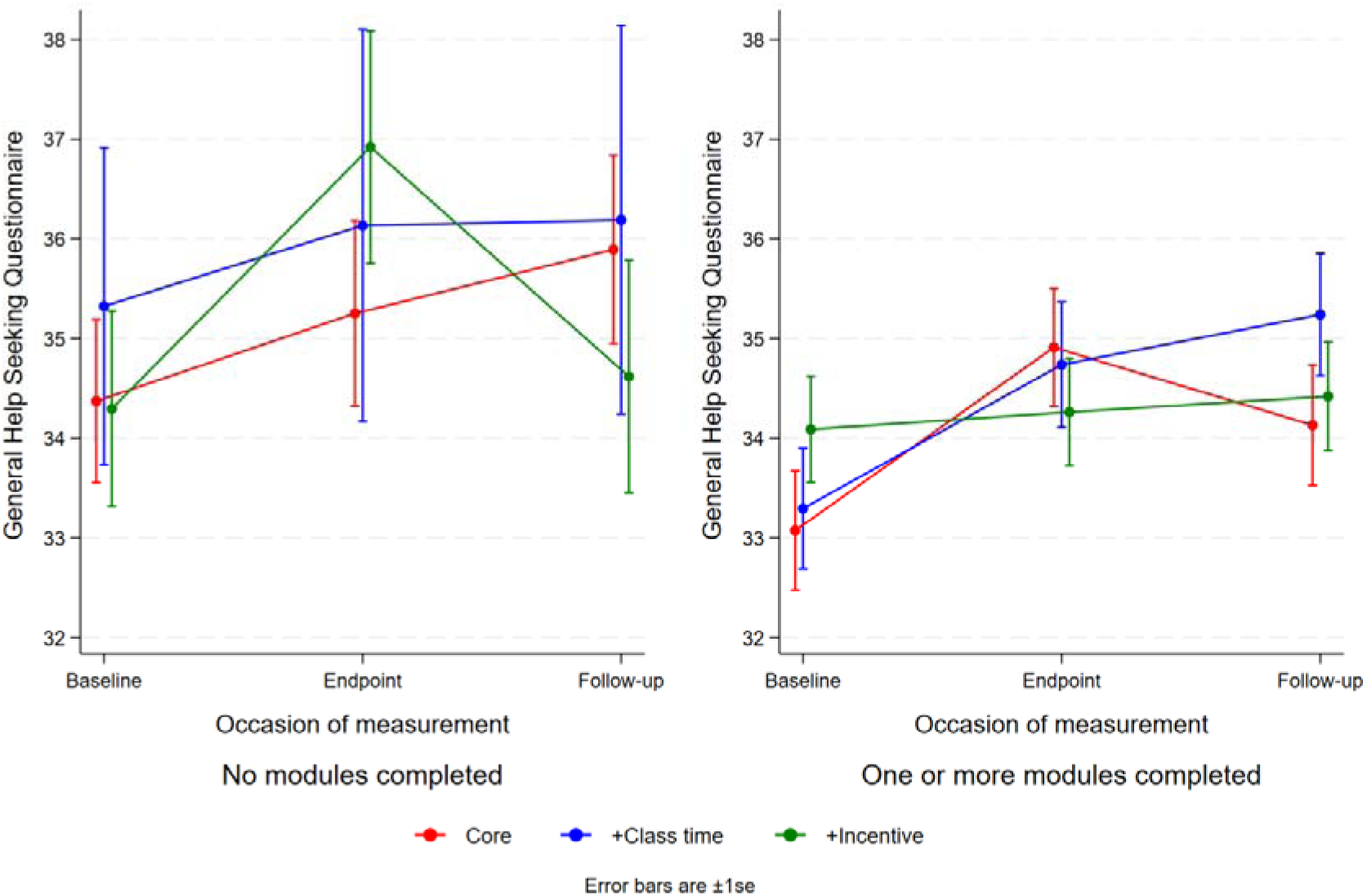
Help-seeking intentions by condition and occasion of measurement for students who accessed (a) no modules, and (b) one or more modules.

### Service satisfaction and barriers to use

As shown in Table 4, across all conditions, most students found the service easy to use and understand, and over half found the service interesting, and enjoyable. However, overall enjoyment and ease of use were lower among those allocated to receive additional class time. Students in the financial incentive condition reported higher usefulness and comfort with service requirements such as providing a mobile number compared to those in the standard service and class time groups. Almost two-thirds of all students were comfortable with the school counsellor follow-ups. Across all trial arms, less than half reported that they would use the service again in the future or reported that they found the service helpful in managing their daily lives or their feelings. Students across all conditions cited personal barriers such as forgetfulness, low motivation, and service mismatch with needs as barriers to service use. Additionally, over one-third of students in the class time and financial incentive groups, and just over a quarter of those in the standard service group, identified time constraints as a barrier. Overall, students who received the standard service reported fewer barriers to service use compared to those in the financial incentive or class time conditions. Interestingly, intervention-specific and technical barriers were less prevalent than personal barriers across all conditions, with a quarter or fewer students citing these issues as impeding their use of the service. Mean helpfulness ratings were 3.21 (SD: 1.00), 2.89 (SD: 1.03), and 3.19 (SD: 0.99) for the standard service, class time, and financial incentive conditions respectively (range: 1 to 5).

**Table 4.**
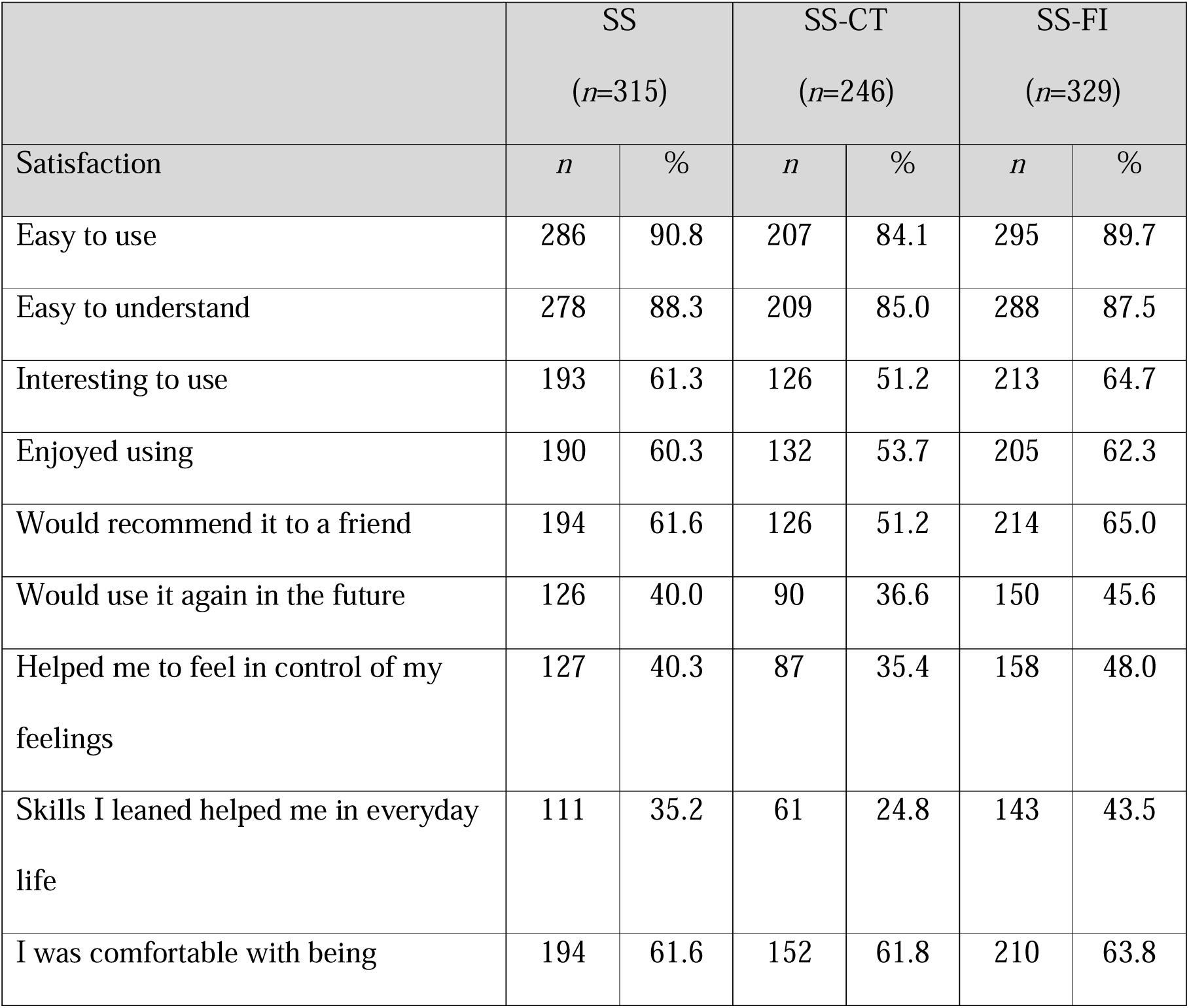
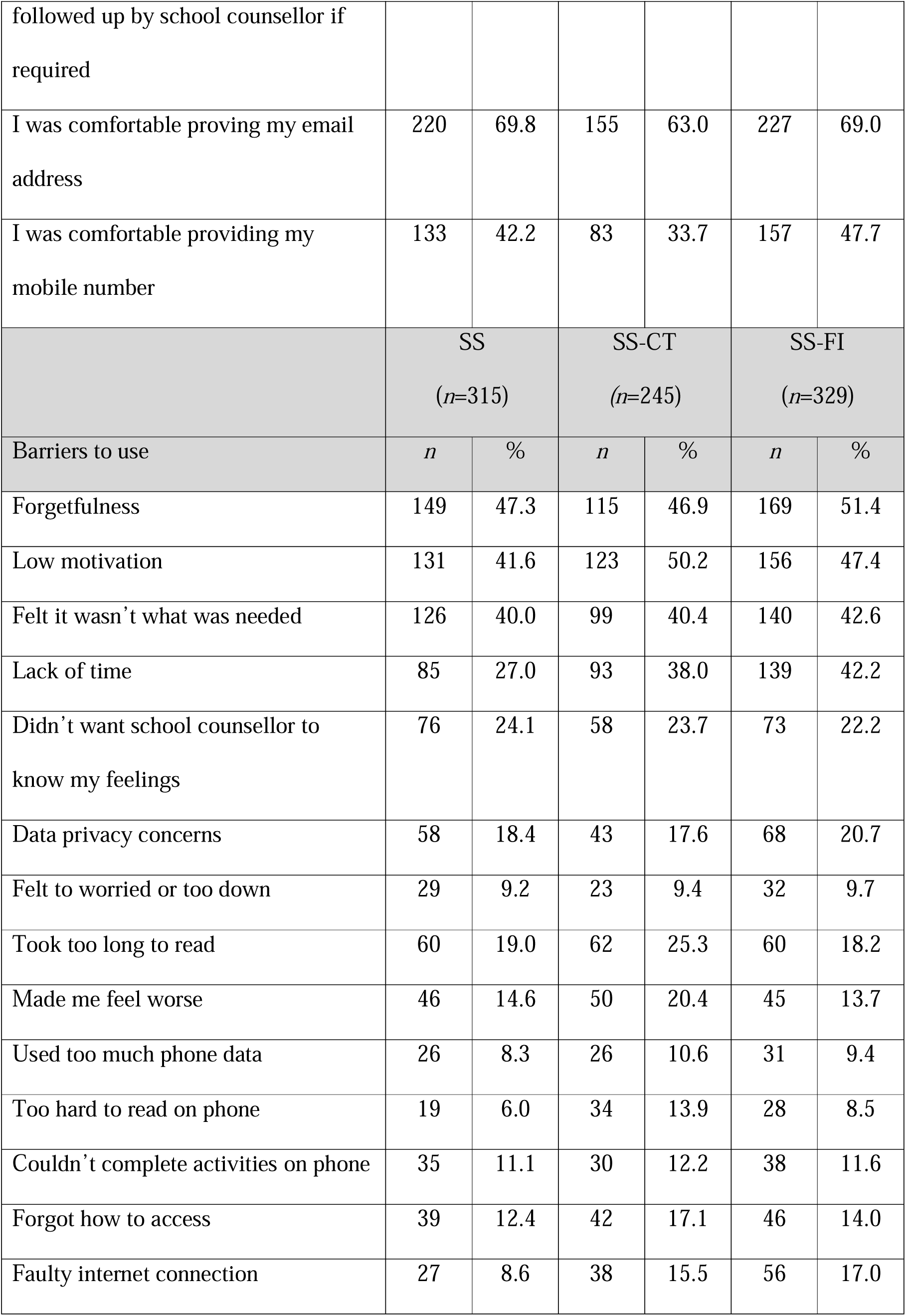
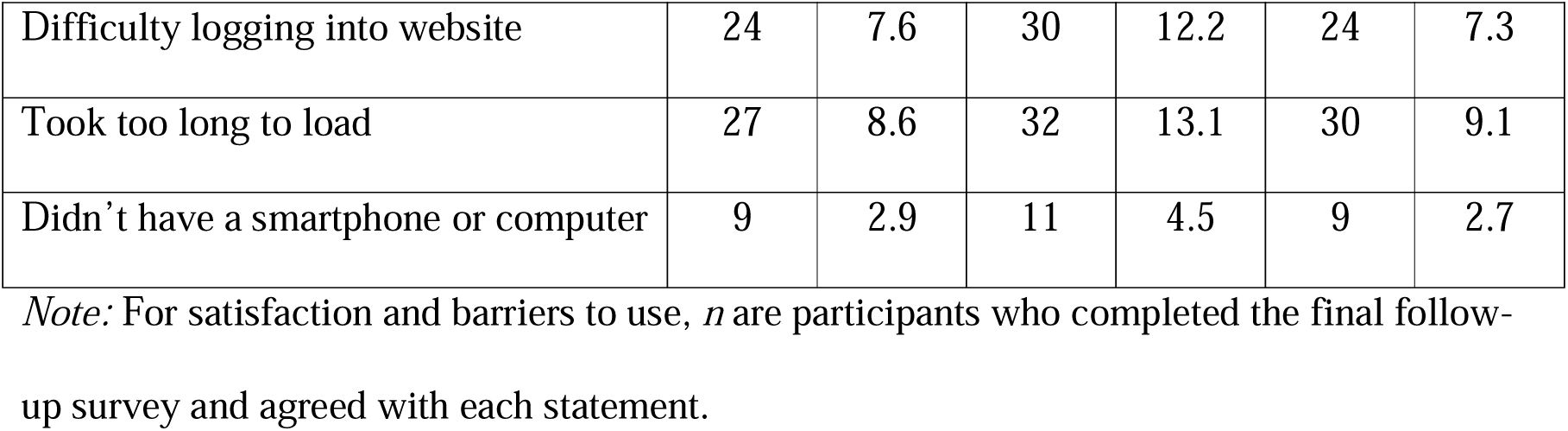
Service satisfaction and barriers to use at 12 weeks post-baseline.

|  | SS<br>(*n*=315) |  | SS-CT<br>(*n*=246) |  | SS-FI<br>(*n*=329) |  |
| --- | --- | --- | --- | --- | --- | --- |
| Satisfaction | *n* | % | *n* | % | *n* | % |
| Easy to use | 286 | 90.8 | 207 | 84.1 | 295 | 89.7 |
| Easy to understand | 278 | 88.3 | 209 | 85.0 | 288 | 87.5 |
| Interesting to use | 193 | 61.3 | 126 | 51.2 | 213 | 64.7 |
| Enjoyed using | 190 | 60.3 | 132 | 53.7 | 205 | 62.3 |
| Would recommend it to a friend | 194 | 61.6 | 126 | 51.2 | 214 | 65.0 |
| Would use it again in the future | 126 | 40.0 | 90 | 36.6 | 150 | 45.6 |
| Helped me to feel in control of my feelings | 127 | 40.3 | 87 | 35.4 | 158 | 48.0 |
| Skills I learned helped me in everyday life | 111 | 35.2 | 61 | 24.8 | 143 | 43.5 |
| I was comfortable with being | 194 | 61.6 | 152 | 61.8 | 210 | 63.8 |
| followed up by school counsellor if required |  |  |  |  |  |  |
| I was comfortable providing my email address | 220 | 69.8 | 155 | 63.0 | 227 | 69.0 |
| I was comfortable providing my mobile number | 133 | 42.2 | 83 | 33.7 | 157 | 47.7 |
|  | SS<br>(n=315) |  | SS-CT<br>(n=245) |  | SS-FI<br>(n=329) |  |
| Barriers to use | *n* | % | *n* | % | *n* | % |
| Forgetfulness | 149 | 47.3 | 115 | 46.9 | 169 | 51.4 |
| Low motivation | 131 | 41.6 | 123 | 50.2 | 156 | 47.4 |
| Felt it wasn't what was needed | 126 | 40.0 | 99 | 40.4 | 140 | 42.6 |
| Lack of time | 85 | 27.0 | 93 | 38.0 | 139 | 42.2 |
| Didn't want school counsellor to know my feelings | 76 | 24.1 | 58 | 23.7 | 73 | 22.2 |
| Data privacy concerns | 58 | 18.4 | 43 | 17.6 | 68 | 20.7 |
| Felt too worried or too down | 29 | 9.2 | 23 | 9.4 | 32 | 9.7 |
| Took too long to read | 60 | 19.0 | 62 | 25.3 | 60 | 18.2 |
| Made me feel worse | 46 | 14.6 | 50 | 20.4 | 45 | 13.7 |
| Used too much phone data | 26 | 8.3 | 26 | 10.6 | 31 | 9.4 |
| Too hard to read on phone | 19 | 6.0 | 34 | 13.9 | 28 | 8.5 |
| Couldn't complete activities on phone | 35 | 11.1 | 30 | 12.2 | 38 | 11.6 |
| Forgot how to access | 39 | 12.4 | 42 | 17.1 | 46 | 14.0 |
| Faulty internet connection | 27 | 8.6 | 38 | 15.5 | 56 | 17.0 |
| Difficulty logging into website | 24 | 7.6 | 30 | 12.2 | 24 | 7.3 |
| Took too long to load | 27 | 8.6 | 32 | 13.1 | 30 | 9.1 |
| Didn't have a smartphone or computer | 9 | 2.9 | 11 | 4.5 | 9 | 2.7 |

*Note:* For satisfaction and barriers to use, *n* are participants who completed the final follow-up survey and agreed with each statement.

## DISCUSSION

This trial represents one of the few attempts to formally evaluate the effectiveness of implementation strategies for increasing student engagement and uptake of a universal school-based digital mental health intervention. Overall, student engagement in the intervention remained low across all three conditions, despite the use of targeted implementation strategies. The evaluated strategies did not lead to significant differences in uptake or retention among students. Although prior research has demonstrated some evidence supporting the use of these strategies for improving individuals’ engagement with interventions in structured settings (e.g., Neil et al., 2009) and increasing retention and participation (e.g., Musiat et al., 2022; Pratap et al., 2020; Wurst et al., 2020), the use of these strategies for supporting the implementation of digital mental health interventions in Australian secondary schools requires further refinement.

Contrary to the hypotheses, students who received financial incentives were not more likely to engage with the service. Students who received financial incentives also showed lower improvement in help-seeking intentions compared to students who received the standard service or extra class time, particularly if they did not complete any modules. In this trial, the use of external rewards may have undermined intrinsic motivation, particularly once the incentive was removed (Deci et al., 1999). The literature emphasises the importance of intrinsic motivation for sustained engagement in interventions (Ryan & Deci, 2000), and highlights the challenges of relying on extrinsic motivators, like financial rewards, for lasting behaviour change (Kwasnicka et al., 2016). For example, a recent study examining the impact of motivation on self-guided, 8-week web-based intervention found a positive correlation between internal motivation and service initiation, intervention adherence, and intervention satisfaction (Hanano et al., 2024). Students with moderate to high internal motivation also exhibited greater symptom improvement. Thus, future research could explore strategies to enhance intrinsic motivation by incorporating elements that foster autonomy (e.g. personalisation of goals and activities, flexible scheduling), and competence (e.g., skill-building exercise, progress tracking) to improve engagement and outcomes in web-based interventions.

Service quality improves when program goals, rationale, and components are clearly communicated, feedback on program outcomes is provided, implementation barriers are addressed with well-developed plans, and individual responsibilities are well-defined (Greenberg & Abenavoli, 2017; Thabrew et al., 2023). Although participating schools were provided with a comprehensive implementation guide, which detailed the service’s purpose and benefits, outlined a step-by-step process for delivery, and was supported with regular emails, telephone calls, and checklists, module access varied widely across schools. Some schools had high levels of engagement, while others had many students who did not access any modules. This variation suggests that factors such as school type, environment, and staff involvement may have influenced engagement. The participating schools ranged from government and non-government to special assistance schools, the latter offering flexible learning environments for students struggling in traditional settings. Schools were also situated in rural, regional, and metropolitan locations, with each context affecting factors such as class structure, attendance, and teacher support. Government schools, for example, generally have a higher student-to-staff ratio compared to non-government schools which include Catholic and independent schools, while rural government schools often face additional challenges such as staff shortages, higher turnover, and limited access to mental health resources like counsellors and psychologists (ACARA, 2022, 2023). Future research involving a larger number of schools could explore whether certain school attributes are associated with higher engagement. Moreover, to improve engagement, tailoring implementation strategies to the unique context of each school and the specific roles of its staff (e.g., Beames et al., 2021; Lyon & Bruns, 2019; Richter et al., 2022) may be more effective than focusing solely on student-level strategies.

Importantly, the COVID-19 pandemic introduced significant challenges, with teachers and schools facing additional disruptions and stressors during this time. During the pandemic, teachers reported heightened stress, exhaustion, and low positivity in their work due to managing the demands of remote learning, combined with increased responsibilities from students, parents, and school leaders (Billett et al., 2023). Many schools in this study shifted the delivery of the service from in-person to online to comply with government guidelines and school preferences, which affected how the service was implemented. During these virtual sessions, a researcher was present on-screen, but student environments varied—some students were placed in smaller classrooms, while others were seated in larger halls or theatres. Remote, online delivery worked well when staff were organised, provided adequate supervision, and actively engaged students. However, module completion rates were likely impacted by inconsistent or low supervision, less engaged staff, and varying compliance with the service requirements in less structured settings.

Additionally, many students may have been overwhelmed by competing demands and stressors during the pandemic, contributing to the low engagement with the modules. Nearly half of the students across all conditions cited forgetfulness, low motivation, and a perception that the service was not what they needed as the primary barriers to using the service. These barriers, such as time constraints and low motivation, are consistent with findings from earlier evaluations of the service (O’Dea et al., 2020) and other similar studies (e.g., Baldofski et al., 2024; Werner-Seidler et al., 2017). It is possible that the additional measures, such as providing a short information video about the service and incorporating more comprehensive feedback following screening, were insufficient to engage students meaningfully. Alternatively, students may not have found the service relevant at the time, particularly in the context of heightened loneliness, isolation, and uncertainty during the pandemic (e.g., Li et al., 2022). While the service offered some degree of personalisation and choice, factors known to boost motivation and engagement with digital mental health interventions (Saleem et al., 2021), future studies could explore alternative or complementary strategies such as moderated peer-support groups or platforms for students to share experiences (e.g., Badawi et al., 2023; Carolan et al., 2017) or incorporate approaches that focus more directly on fostering intrinsic motivation (e.g., Hanano et al., 2024).

### Limitations

The findings should be viewed considering several limitations. First, the impact of COVID-19 on data collection may have influenced student engagement, as disruptions to normal school operations and pandemic-related stressors, could confound the study results. Second, the final sample consisted of students from 20 out of 119 interested schools, with two schools withdrawing due to the pandemic. This non-random attrition introduces potential selection bias which could affect the generalisability of the results. Geographical and contextual limitations also apply, as the sample primarily included schools from NSW and one independent school in SA, limiting the generalisability to schools in other states or territories in Australia or countries with different cultural or educational contexts. Additionally, participation was contingent upon obtaining support from school principals and approval from governing ethical bodies, excluding schools in certain Catholic Dioceses within NSW and outside of NSW. The reliance on principal support could have excluded interested staff members who were unable to secure approval for the service in their schools. Further, effective evidenced-based strategies at the school level include monitoring progress, providing feedback to the implementation team, securing teacher buy-in, and organising regular meetings with school personnel (Baffsky et al., 2023; Beames et al., 2023). Although staff received regular emails, calls, and checklists to facilitate communication, review processes, and track progress, the pandemic and the scheduled disruptions led to a shift in resources toward rescheduling school visits, developing additional protocols, and supporting schools for online service delivery. As a result, we did not formally collect data on adherence to the implementation protocol due to feasibility constraints. This lack of adherence data may limit our understanding of how consistently the program was delivered across schools. Ensuring and safeguarding program fidelity should be a key focus for future research.

## Conclusion

The strategies of providing extra class time allocation and financial incentives did not significantly improve student engagement or help-seeking outcomes. Variability in school environments, staff involvement, and external stressors—particularly those related to the COVID-19 pandemic—likely contributed to the inconsistent levels of engagement across schools. These findings highlight the importance of considering the broader school context when implementing mental health services. Future efforts should prioritise strategies that are flexible and targeted towards schools to enhance student participation in digital mental health programs to improve student outcomes.

## Data Availability

All data produced in the present work are contained in the manuscript

## Declaration of funding and/or competing interests

The Smooth Sailing service was developed by researchers at the Black Dog Institute. This study was supported by The Prevention Hub, Australian Government Department of Health.

## Author contributions

BOD, MSK, and MRA designed and built the service. BOD and MSK conceived this project and BOD, MSK, HC, and NC were awarded the funding. BOD, MSK, SB, BP, and MRA designed the study protocol. MSK, DG, BP, and MRA coordinated trial recruitment and trial operations and SB and CC assisted with research support including the school visits. AJM was the trial statistician who conducted all the analyses. MSK and BOD prepared the manuscript, and all authors reviewed the final manuscript.

## Acknowledgments

The authors would like to acknowledge the involvement of the students and school staff in the trial. The authors would also like to acknowledge the Trial Steering Group including Professor Jill Newby, Associate Professor Alexis E. Whitton, Professor Fiona Shand, Professor Kathryn Boydell, Professor Vijaya Manicavasagar, Dr Rebecca Hardy, Kate Cope, and Nicole Cockayne. The authors also acknowledge Amy O’Halloran and Hiroko Fujimoto for providing administrative support for several school visits.

