## Supplementary File for "Optimising the implementation of a universal web-based mental health service for Australian secondary schools: A cluster randomised controlled trial"

**Supplementary Material**

**Table 1.** ERIC Implementation strategies integrated into the Smooth Sailing service

| **Target** | **Strategy** | **Activity** |
| --- | --- | --- |
| **School** | **Audit & Provide feedback**: Collect and summarise data regarding implementation of the new program over a specified period and give it to administrators and school personnel to monitor, evaluate, and support implementer behaviour. | Each school received a school report upon completion of the service that outlined their students’ mental health and peer-norm comparisons. |
|  | **Centralise technical assistance:** Develop and use a centralised system to deliver and facilitate access to technical assistance focused on implementation issues. | The service team assumed responsibility for all IT requirements including facilitating school access to the service URL and directly responding to all IT inquiries. |
|  | **Develop a detailed implementation plan**: A blueprint that includes the intended goals to be achieved alongside processes and strategies to achieve those goals. | An electronic and paper-based Implementation Guide was developed in consultation with key school staff. It outlined the roles and responsibilities of service personnel and was distributed to all schools prior to implementation. |
|  | **Promote adaptability:** Identify the ways the program can be tailored or adapted to best fit with the school/classroom context, meet local needs, and clarify which elements of the new practice must be maintained to preserve fidelity. | The implementation team worked with each school to devise a plan for how the service could be implemented. For example, delivered to a whole year group at once or in smaller class sizes, in different free class periods, at times with fewer competing priorities. |
|  | **Identify and prepare champions:** Individuals who dedicate themselves to supporting, marketing, and driving through an implementation, overcoming indifference or resistance that the intervention may provoke in a school or district. | The implementation team identified and formally recognised the school-based champions. These champions were promoted as the Implementation Team and trained accordingly. |
|  | **Organise school personnel implementation team meetings***:* Develop and support teams of school personnel who are implementing new practices and give them protected time to reflect on the implementation effort, share lessons learned, and support one another’s learning. | Contact between the implementation team and the school was facilitated by a “school liaison” who was an implementation team member who managed each of the schools’ requirements. The school liaisons also visited the schools to conduct the service delivery in-person, providing on the ground support to the school staff. |
|  | **Visit other sites:** Visit sites where a similar implementation effort has been considered successful. | All school liaisons visited various sites to observe successful implementation sessions across schools. |
|  | **Conduct educational outreach visits and provide ongoing coaching:** Have a trained person meet with school personnel in their practice settings to educate them about new practices with the intent of changing the school personnel’s practice. | Upon completion of each screening session, school liaison staff spent time with the school counsellor to review the student follow-ups, discuss their plan, and ensure they felt appropriately supported. |
|  | **Develop educational materials:** Develop and format manuals, toolkits, and other supporting materials in ways that make it easier for stakeholders to learn about new practices and for school personnel to learn how to deliver the new practices with fidelity. | The service team collated and disseminated various educational materials to support the service components including manuals, frequently asked questions, sample verbal scripts, that were updated as the service implementation progressed. |
|  | **Remind school personnel:** Develop reminder systems (e.g., email prompts or visual cues) designed to help school personnel recall information and/or prompt them to deliver core components of new practices. | The service team followed a communication plan for each of the schools to build rapport, remind them of key dates, and to problem solve any implementation challenges. |
| **Student** | **Prepare families and students to be active** **participants** Prepare families and/or students to create “pull” (i.e., motivation or pressure to implement) for the delivery of the new practice by asking relevant questions, advocating for the new practice, and inquiring about guidelines for implementation, the evidence and rationale behind decisions, or about other effective new practices that could be implemented. | Developed and integrated a student video explainer for the service to be disseminated to all students prior to service delivery.  Developed and integrated an online consent procedure that enabled students to give their consent online.  Communication materials for parents were distributed via the schools using usual methods of communication. |
|  | **Alter and provide individual- and system-level incentives:** Work to provide individual- (e.g., recognition and acknowledge, gift card) and/or system-level incentives to districts or schools to participate (e.g., grant money, free training, and consultative support) and engage in an implementation effort involving a new practice. | Financial incentives in the form of a gift card were embedded to encourage students to complete the service activities. |
|  | **Alter student or school personnel obligations to enhance participation in or delivery of new practice, respectively:** Create structures where students or school personnel are relieved of a particular obligation for participating in or delivering more preferred practices/supports than less-preferred practices/supports. | Components of the service were adapted to align with the school curriculum to allow schools to allocate class time to the activities. |

**Intervention: The Smooth Sailing Service.**

The Smooth Sailing Service is a 12-week, web-based mental health program designed to support youth mental health through structured, stepped care. The service uses an online platform to register students, conduct initial and follow-up mental health screenings, and allocate students to an appropriate step of care. Smooth Sailing provides students with evidence-based information, self-guided mental health activities, and referrals to a school counsellor when needed.

The service is delivered over three classroom sessions spaced six weeks apart, allowing students to explore two main program components: the Bite Back online mental fitness challenge during the first six weeks, and a series of psychoeducational modules in the second six weeks. Members of the research team and school staff (teachers, year advisors and/or school counsellors) are present during these sessions to address questions, provide guidance, and offer assistance as needed.

1. **Registration and initial screening.**

During the first classroom session (i.e., the baseline assessment and initial screening session), students completed registration and screening questions using a compatible device (e.g. desktop computer, laptop, tablet, or smartphone). To register, students were asked to visit the Smooth Sailing website using a school-specific URL, complete an online consent form, provide a valid, active, and accessible school-administered email address (or personal email address if the school email address was unavailable), and create a password. Students then completed the initial mental health screening using the 2-item Generalised Anxiety Disorder Scale (GAD-2; Spitzer et al., 2006) and the 2-item Patient Health Questionnaire (PHQ-2; Kroenke et al., 2003). If symptom thresholds were met, the measures expanded to the GAD-7 and PHQ-8 (Kroenke et al., 2001), with responses scored the same as the full-scale (Löwe et al., 2010). Using the total scores of the GAD and PHQ, the service automatically allocated students to one of three steps of care (see Figure 1) and provided a personalised online dashboard with recommended activities. Although students were not informed of their step allocation, they received feedback on their score range and an explanation of how the suggested activities are beneficial. There were two versions of the score range and rationale for engaging with activities: one for students at Step 1 and another for students allocated to Steps 2+.

2. **Follow-up assessments and step allocation.**

At 6-weeks (i.e., the second screening assessment) and 12-weeks (i.e., the third and final screening assessment; primary endpoint), students were sent a URL to their registered email to complete the GAD and PHQ screening process again. An algorithm re-calculated students’ step based on the level of anxiety and depressive symptoms indicated by the GAD and PHQ completed at baseline and at the 6-week time point. Students were automatically allocated to one of five symptom severity categories and one of three steps of care following the *Australian Practice Guidelines for Clinical Depression* (McDermott et al., 2010). The level of care, or intensity of care, is matched to the level (i.e., severity) of symptoms and increases cumulatively (See Figure 1). Students received a personalised dashboard that displayed modules corresponding with their six-week step allocation and feedback on their mental health score range. The dashboard also provided an overview of the psychoeducation modules and their recommended activity. The self-directed web-based psychoeducation provided youth-oriented information about mental health, depression, anxiety, and help-seeking (see Table 2). An additional sixth module provided information about a selection of publicly available, free, evidence-based online CBT programs (see Table 3).

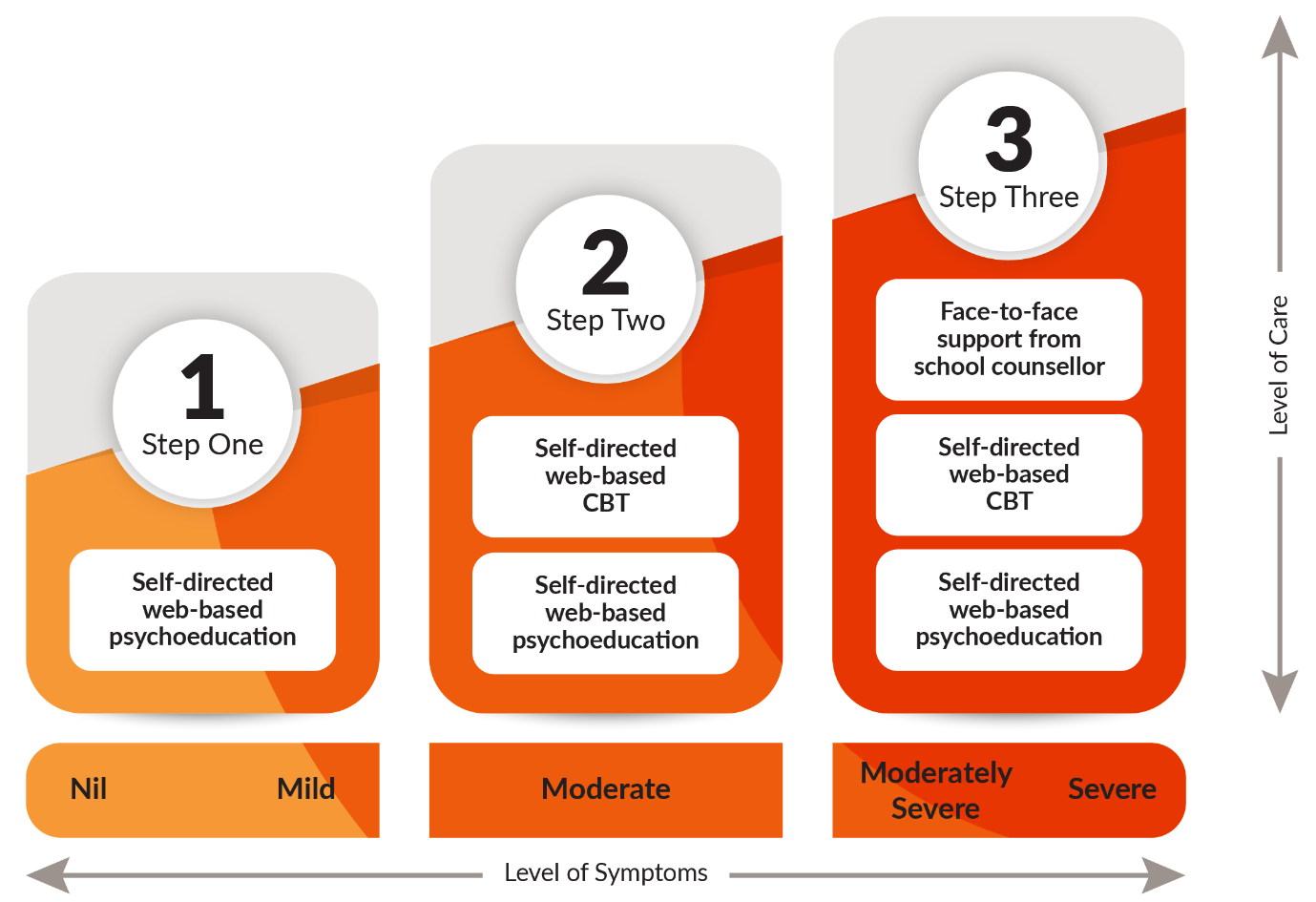

**Figure 1. The Smooth Sailing service uses a stepped care approach such that the level of care is matched to the level of symptoms and increases cumulatively.**

The step assessment was repeated at 12-weeks (i.e., the third and final screening time point) which reallocates the level of care, depending on the student’s results. If a student has not responded (i.e., symptoms remain elevated or have worsened) within six weeks, they are stepped up to the next level of care. The combined step allocation accounts for the student’s previous step, their current step, and whether there has been any change. There is no stepping down in the current model. As such, the level of care remains the same and students were instructed to continue using the service as advised by the personalised dashboard without losing access to any of the previously accessible content, programs, and apps.

3. **Step-specific Interventions.**

There were three versions of the score range and rationale for engaging with activities that corresponded to students allocated to Steps 1, 2, and 3.

**Step One. Self-directed web-based psychoeducation.** Students with nil to mild symptoms (i.e., symptom severity categories “0” and “1”) were allocated to Step One. For students at step 1, the service offered five 10-minute self-directed interactive modules on general mental health, anxiety, depression, and help-seeking that were complimented by animations, illustrations, and hyperlinks to credible youth mental health services and websites (see Table 2). All module content was created specifically for the Smooth Sailing service and was reviewed by youth and health professionals in the co-design process. The content was also edited by a copywriter to ensure readability for young adolescents.

**Step Two. Self-directed web-based CBT.** Students with moderate symptoms (i.e., symptom severity category “2”) were allocated to Step Two. In addition to having access to the five psychoeducation modules, the dashboard included an additional sixth help-seeking module, which was displayed first. This module referred students to three publicly available, free, evidence-based, self-directed Internet Cognitive Behaviour Therapy (iCBT) interventions provided by Australian mental health organisations and universities: (i) WeClick (O'Dea et al., 2020); (ii) MoodGym (Calear et al., 2009) (iii) The BRAVE Program (Spence et al., 2011) (see Table 3). Because the latter two programs were offered by external providers, the research team did not have access to the uptake and data collected by these.

**Step Three. Face-to-face support from a school counsellor.** Students with moderately severe to severe symptoms (i.e., symptom severity categories “3” and “4”) and/or reported ‘thoughts that that they would be better off dead or of hurting themselves’ (i.e., score > 0 on item 9 of the PHQ-9) were allocated to Step Three and, in addition to having access to all six Smooth Sailing modules, triggered a follow-up notification with a school counsellor for face-to-face support. School counsellors tracked these notifications using a purpose-built secure web portal. For any student who reports having thoughts of self-harm or death in the past two weeks, a more in-depth follow-up assessment was conducted by the school counsellors during school time but after completion of the school visit to determine their true risk status and whether contact had to be initiated with the student’s parent(s)/guardian(s), a mandatory report made, or external referral required. School counsellors follow their normal school protocols and duty of care procedures when attending to these student referrals.

**Table 2.** Title and Description of Smooth Sailing Modules.

| **Module** | **Title** | **Description** |
| --- | --- | --- |
| **1** | What is mental health? | Information about mental health issues that are common among young people and when it might be time to seek help. |
| **2** | Feeling on edge | Information about anxiety, how to identify it, potential causes, how and where to seek help, and practical tips for managing it. |
| **3** | Waves of sadness | Information about depression, differences between sadness and depression, potential causes, how and where to seek help, and practical tips to cope. |
| **4** | When it’s time to tell someone | Information about when to seek help, how to talk to friends and parents/carers, seeking help from a General Practitioner, and the roles of different health professionals. |
| **5** | When a mate needs a hand | Information about ways to help others including having a private chat and seeking help together, respecting the treatment process, and the importance of looking after yourself. |
| **6^a^** | Don’t fret, help is here | Information about a selection of publicly available, free, evidence-based online CBT programs. |
| ^a^Available to students allocated to Step Two or Step Three but not Step One. | | |

**Table 2.** Evidence-based Online CBT Programs Recommended by Smooth Sailing.

| **Program**^a^ | **Description to students** |
| --- | --- |
| **MoodGym** | An interactive program that will help you identify what makes you upset and provide strategies to help tackle those emotions and change the way you think. |
| **The BRAVE** | An online activity that can lower feelings of worry in young people. It will help you learn how to identify anxiety and stress, develop relaxation skills, and replace negative thinking with more positive thinking. |
| **WeClick** | A relationships-focused program that will help you learn skills and strategies to improve your friendships and other relationships. |
| ^a^Available to students allocated to Step Two or Step Three, but not Step One. | |

**Safety protocols and stepped care model.** Following the safety protocol, any student who had not responded to their allocated care (i.e., symptoms remained elevated or had worsened) at 6-or 12-weeks was stepped up to the next level of care. Due to the novelty of the service model and the short study period, no students were stepped down. As such, the level of care remained the same and students were instructed to continue using the service as advised by the personalised dashboard without losing any access to any of the previously accessible content, programs, or apps. Access to the service ceased two weeks after the third and final school visit.

**Monitoring.** Following the 6-week assessment, students allocated to Step 3 were invited to participate in fortnightly mood check-ins delivered via short message services (SMS) or email. These check-ins included the GAD-2 and PHQ-2 symptom scales to monitor anxiety and depressive symptoms. A total of four check-ins were conducted during the study period, each accompanied by two reminder messages to encourage completion. Students received generalised feedback based on their responses and were prompted to continue engaging with the service.

**Strategies to improve adherence to intervention protocols.**

1. **Reminders.** All participating students received three reminders to complete the Bite Back Mental Fitness Challenge, as well as an additional three reminders to complete the Smooth Sailing modules. Students allocated to receive a financial incentive had additional information about the incentive included in the three Smooth Sailing module reminders.
2. **Class time for registration and screening.** Schools provided dedicated class time for students to complete registration and screening activities, facilitating initial engagement with the intervention. After completing the screening questionnaires, students were given time to explore both Bite Back and the Smooth Sailing modules.
3. **Verbal reminders and encouragement.** School staff and research team representatives offered verbal reminders and encouragement during the screening sessions. School staff and research team representatives encouraged participating students to engage with Bite Back and the Smooth Sailing modules for the six weeks respectively that each program was accessible and active.
